# Intellectual Property Literacy, Innovation Readiness and Innovation Practice in Syria’s Pharmaceutical Sector: A Cross-Sectional Study

**DOI:** 10.64898/2026.06.22.26356119

**Authors:** Chadi Khatib, Hala Alkozy, Zainab Hamdan, May Isber, Jawa Mlhem

**Affiliations:** Faculty of Pharmacy, Manara University, Syria

**Keywords:** intellectual property literacy, innovation readiness, pharmaceutical innovation, innovation practice, pharmaceutical sector, Syria, cross-sectional study

## Abstract

**Background:** Innovation in pharmaceutical sectors operating under resource and institutional constraints may depend not only on knowledge and attitudes but also on the conditions that enable innovation-related activities to occur. This study examined the relationships among intellectual property (IP) literacy, innovation attitudes, innovation readiness, and reported innovation practice among pharmaceutical professionals in Syria.

**Methods:** A cross-sectional survey was conducted among 303 pharmaceutical professionals between March and April 2026. Four composite indices were constructed to assess IP literacy, innovation attitudes, innovation readiness, and innovation practice. Descriptive statistics, correlation analyses, group comparisons, and multivariable regression models were used to characterize patterns of association among study domains. The analysis was designed to identify empirical patterns rather than infer causal relationships.

**Results:** Innovation attitudes were comparatively high (73.56/100), whereas innovation readiness (17.00/100) and innovation practice (12.65/100) were substantially lower. IP literacy was positively associated with innovation readiness (r = 0.384, p < 0.001) and innovation practice (r = 0.205, p < 0.001). In contrast, innovation attitudes were not significantly associated with reported innovation practice (p = 0.332). Regression analyses indicated that the inclusion of innovation readiness improved model fit beyond specifications based on knowledge and attitudes alone (ΔR² = 0.058, p = 0.028). Significant differences in readiness and practice were observed across professional groups (p < 0.001), whereas knowledge and attitudes showed limited variation.

**Conclusions:** High levels of innovation-related knowledge and positive attitudes did not correspond to high levels of reported innovation practice in this setting. The findings suggest that innovation readiness may capture enabling conditions that are not reflected by knowledge or attitudinal measures alone. These results support the value of examining contextual and institutional factors when assessing innovation capacity in resource-constrained pharmaceutical systems. Given the substantial gap observed between innovation attitudes and innovation practice, educational strategies may represent one avenue for strengthening innovation readiness. In the Syrian context, strengthening innovation-oriented education and university–industry engagement may help cultivate innovation competencies and support the translation of research into practical applications.

## 1. Introduction

Pharmaceutical innovation in transitional and resource-constrained economies is shaped not only by the availability of knowledge but also by the institutional conditions that determine whether knowledge can be translated into practice. In pharmaceutical settings, intellectual property (IP) literacy is widely recognized as an important component of innovation capacity because it influences incentives to invest in, protect, and commercialize new ideas. However, evidence from pharmaceutical policy and innovation research suggests that knowledge alone does not adequately explain differences in innovation performance across systems, particularly the persistent gap between innovation potential and realized innovation activity (Cockburn, 2009; Birkbeck, 2016; OECD, 2018; Gamba, 2017; Lee et al., 2021).

A substantial body of applied research has relied on the Knowledge–Attitude–Practice (KAP) framework, which assumes that knowledge promotes favorable attitudes and that these attitudes subsequently influence practice. Although this framework remains widely used, empirical findings increasingly suggest that the relationship between knowledge, attitudes, and behavior is neither uniform nor necessarily linear. Recent studies have employed mediation and structural modelling approaches to examine these relationships (Cheng et al., 2024; Qin et al., 2024), yet the extent to which such patterns generalize across institutional settings remains uncertain.

Evidence from public health and related fields has repeatedly shown that knowledge and attitudes may be weakly associated with actual behavior when individuals operate under structural constraints, regulatory uncertainty, or limited institutional support (Liao et al., 2022; Luo et al., 2022; Sujarwoto et al., 2022; Wang et al., 2023; Cheng et al., 2024; Qin et al., 2024). These findings suggest that innovation-related behavior may depend not only on what individuals know or believe, but also on whether the surrounding environment enables innovation-related activities to occur.

This observation directs attention toward contextual conditions that may facilitate or constrain innovation. In transitional pharmaceutical systems, such conditions may include access to innovation infrastructure, exposure to applied research environments, organizational support, and the broader regulatory environment. Within the innovation readiness literature, these factors are increasingly conceptualized as enabling conditions that influence whether innovation can be implemented rather than merely intended (Chen et al., 2014; Van den Hoed et al., 2022). Similarly, research on pharmaceutical innovation in middle-income settings highlights the importance of institutional and market conditions in shaping innovation activity (Alzarea et al., 2022; OECD, 2018; Arman et al., 2021).

Despite growing interest in intellectual property, innovation capacity, and pharmaceutical development in low- and middle-income settings, empirical evidence examining IP literacy, innovation attitudes, innovation readiness, and innovation practice within a single analytical framework remains limited. Existing studies have often examined these domains separately, providing limited insight into how they coexist within constrained pharmaceutical systems. This gap is particularly relevant in transitional economies, where institutional constraints may influence the translation of innovation-related capabilities into practice.

In the present study, innovation readiness is conceptualized as a system-level proxy reflecting enabling institutional and structural conditions. It is not treated as an individual psychological trait or latent motivational construct. Rather, it represents reported contextual conditions that may influence the feasibility of innovation-related activity. Accordingly, readiness is examined as a descriptive indicator of the environment within which knowledge and attitudes may or may not be reflected in reported innovation practice. The conceptual framework guiding the study is presented in Figure 1.

**Figure 1.**
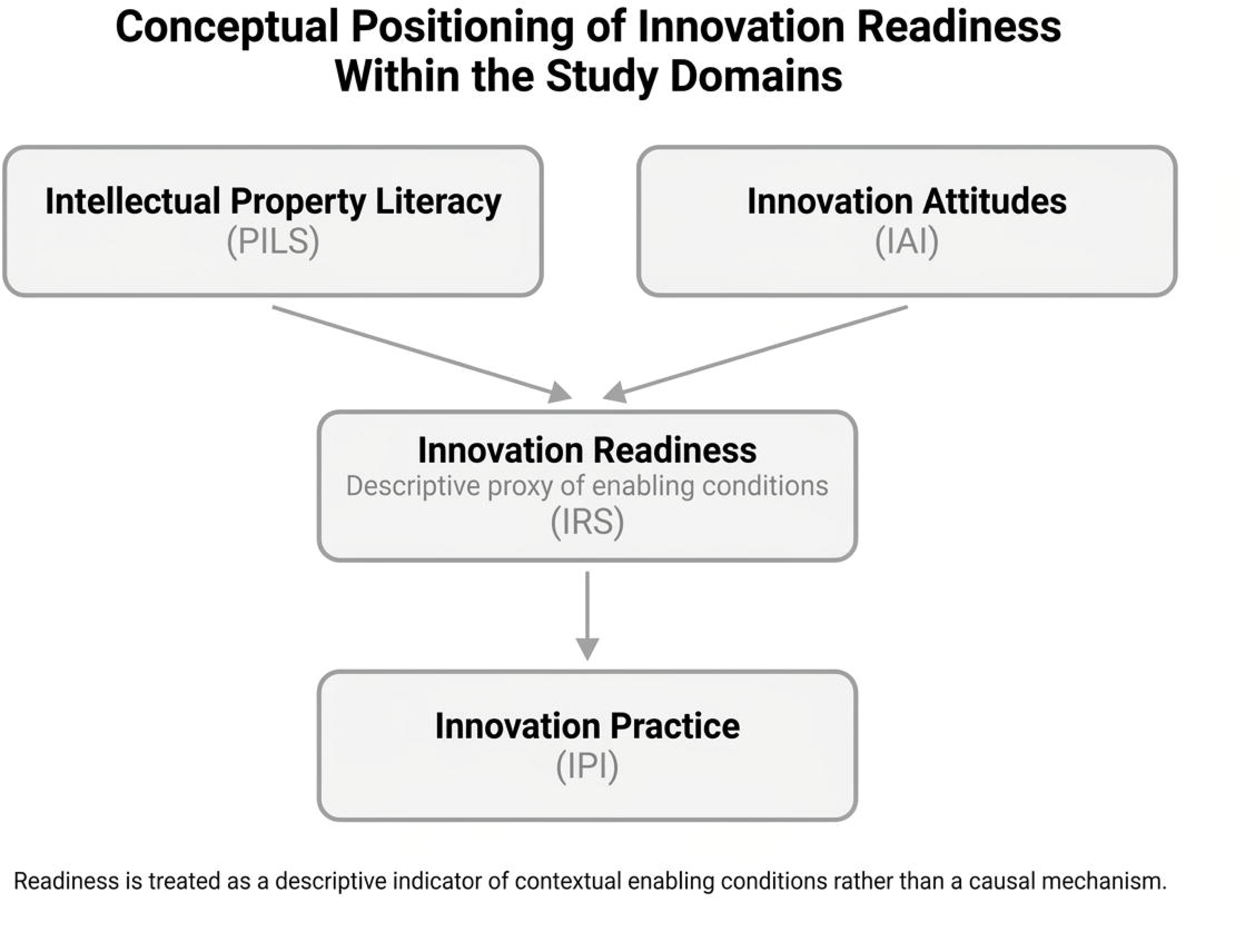
Conceptual Positioning of Innovation Readiness within the Study Domains.

The pharmaceutical sector in transitional economies provides a particularly relevant setting for examining these relationships because it often combines strong normative support for innovation with limited implementation capacity. This situation reflects broader discussions of innovation traps and catch-up constraints, where favorable orientations toward innovation do not necessarily translate into innovation outcomes (Arman et al., 2021; Lee et al., 2021; OECD, 2018). Consequently, the sector offers a useful context for exploring whether observed differences between knowledge, attitudes, readiness, and practice are more strongly associated with institutional conditions than with individual-level factors alone.

The aim of this study was to examine the relationships among IP literacy, innovation attitudes, innovation readiness, and reported innovation practice in Syria’s pharmaceutical sector. Specifically, the study assessed whether innovation readiness provided additional descriptive information beyond conventional knowledge–attitude approaches. Given the cross-sectional design, the analysis focused on identifying empirical patterns of association rather than testing causal relationships.

## 2. Methods

### 2.1 Study Design and Participants

This cross-sectional observational study examined the relationships among intellectual property (IP) literacy, innovation attitudes, innovation readiness, and reported innovation practice within Syria’s pharmaceutical sector. Data were collected between 10 March and 15 April 2026 using an online questionnaire administered through Google Forms.

The study was conducted in accordance with the ethical principles of the Declaration of Helsinki and was approved by the Research Ethics Committee of Manara University, Syria (Approval No. MU-URO-26-02). Prior to accessing the questionnaire, participants were presented with an electronic informed consent form embedded within the survey platform. The consent form described the study objectives, the voluntary nature of participation, procedures for data handling, and the anonymity of responses. Participants were able to proceed to the questionnaire only after providing electronic informed consent. No personally identifiable information was collected at any stage of the study.

Participants were recruited through professional networks, academic institutions, and pharmaceutical manufacturing facilities. Eligibility was restricted to pharmacy students and pharmacy graduates currently studying or working within the Syrian pharmaceutical sector. A convenience sampling approach was employed because no comprehensive sampling frame was available for the target population. To reduce the likelihood of duplicate participation, survey settings were configured to permit only a single response per participant.A total of 400 invitations were distributed, yielding 303 complete responses (response rate: 75.8%). Eligible participants were required either to hold a pharmacy degree or to be actively enrolled in pharmacy education at the time of the survey. Individuals with non-pharmacy educational backgrounds, duplicate submissions, or questionnaires containing substantial missing data (>50% incomplete responses) were excluded. The final analytical sample comprised 303 participants. Participant recruitment and sample derivation are summarized in Supplementary Figure S1.

Internal consistency coefficients are reported for transparency and comparability with previous KAP-oriented studies but were not used as criteria for indicator selection or scale refinement.

### 2.2 Data Collection and Analytical Orientation

The study was designed as an exploratory observational investigation aimed at describing relationships among IP literacy, innovation attitudes, innovation readiness, and innovation practice within a resource-constrained pharmaceutical setting. Given the cross-sectional design, the analysis focused on identifying empirical patterns of association rather than testing causal relationships or temporal pathways.

Data were collected electronically and underwent screening, coding, and statistical analysis. No preregistered analytical protocol was established prior to data collection. To enhance transparency, a post-analysis documentation record was prepared describing analytical decisions, variable construction procedures, and robustness assessments (Supplementary Document S1).

All study variables were operationalized as observed composite indicators and analyzed at the composite-score level. No latent-variable modelling procedures were applied. Alternative model specifications were compared to evaluate the extent to which different combinations of study domains described observed variation in innovation-related practice. Accordingly, all reported findings should be interpreted as observational associations rather than evidence of causality, mediation, or temporal ordering (Maxwell & Cole, 2007).

Reporting was informed by recommendations for observational KAP-related research, including principles outlined in the ChecKAP reporting framework (Zarei et al., 2024). In addition, the study is reported in accordance with the STROBE Statement for cross-sectional studies, and the completed checklist is provided as supplementary material.

### 2.3 Domains and Indicators

Four domains were assessed: pharmaceutical intellectual property literacy, innovation-related attitudes, innovation readiness, and reported innovation practice. Consistent with formative measurement principles, each domain was constructed as a composite index intended to summarize conceptually distinct but complementary indicators rather than interchangeable manifestations of an underlying latent trait (Coltman et al., 2008; Hanafiah, 2020).

To facilitate comparison across domains, all scores were transformed to a standardized 0–100 scale.

#### Pharmaceutical Intellectual Property Literacy Score (PILS)

PILS summarizes objective intellectual property knowledge using nine binary items (K2–K10) covering key pharmaceutical IP concepts, including patents, trademarks, licensing provisions, and international agreements. Correct responses were coded as 1 and incorrect responses as 0 before normalization to a 0–100 scale. Internal consistency (KR-20 = 0.485) is reported for transparency only.

A separate self-assessment item (Q10) measured perceived knowledge and was reserved exclusively for convergent validity assessment. It was not incorporated into PILS computation.

#### Intellectual Property Attitude Index (IAI)

IAI summarizes respondents’ attitudinal orientations toward intellectual property and pharmaceutical innovation using ten Likert-type items (A1–A10). Responses ranged from 1 (“strongly disagree”) to 5 (“strongly agree”). Negatively phrased items (A2 and A5) were reverse coded prior to score calculation. The composite score was subsequently normalized to a 0–100 scale. Cronbach’s alpha was 0.481.

#### Innovation Readiness Score (IRS)

Innovation readiness was conceptualized as a composite indicator reflecting reported enabling conditions relevant to innovation-related activity. The index was intended to describe contextual conditions and was analyzed separately from knowledge and attitudinal measures.

#### Innovation Practice Index (IPI)

Reported innovation-related behavior was summarized using three binary indicators capturing participation in innovation-oriented activities, including intellectual property applications, innovation projects, and research collaborations. Responses were coded 0/1 and normalized to a 0–100 scale. Cronbach’s alpha was 0.524.

#### Diagnostic Gap Indicator

The diagnostic gap indicator (IAI − IPI) was calculated to provide a descriptive measure of the difference between innovation attitudes and reported innovation practice. This indicator was used for descriptive purposes only and was not interpreted as a causal or latent construct. (Edwards, 2001).

#### Measurement Model Considerations

The indices used in this study were constructed as formative composite indicators designed to summarize conceptually distinct dimensions rather than interchangeable manifestations of a single latent construct. Accordingly, internal consistency coefficients are reported for transparency and comparability with previous studies but were not used as criteria for indicator selection or scale refinement. Under a formative measurement framework, moderate internal consistency does not necessarily indicate inadequate measurement quality because individual indicators are expected to contribute unique information to the composite score (Edwards, 2001; Coltman et al., 2008; Hanafiah, 2020).

### 2.4 Statistical Analysis

Descriptive statistics, including means, standard deviations, and observed ranges, were calculated for all composite indicators.

Differences across professional categories (practicing pharmacists, pharmacy students, and academics) were examined using one-way analysis of variance (ANOVA). Statistically significant omnibus tests were followed by Tukey’s honestly significant difference (HSD) procedure for pairwise comparisons. Effect sizes were reported using eta-squared (η²) and Cohen’s d where appropriate.

Bivariate associations among domains were assessed using Pearson correlation coefficients. Ninety-five percent confidence intervals were calculated using Fisher’s z transformation.

To evaluate alternative descriptive specifications, ordinary least squares regression models with HC3 heteroscedasticity-consistent standard errors were estimated. Specification A examined correlates of innovation readiness, whereas Specification B examined correlates of innovation practice. Standardized regression coefficients (β), partial R² values, and variance inflation factors (VIFs) were reported to facilitate comparative interpretation.

Model diagnostics included the Shapiro–Wilk test for residual normality, the Breusch–Pagan test for heteroscedasticity, and the Durbin–Watson statistic for residual autocorrelation (Barker & Shaw, 2015). Because heteroscedasticity was detected in several specifications, HC3 robust standard errors were retained as the primary inferential estimator.

The comparative contribution of readiness-oriented specifications was evaluated through nested model comparisons using changes in explained variance (ΔR²), Akaike Information Criterion (AIC), and Bayesian Information Criterion (BIC). Statistical significance of ΔR² was assessed using the conventional F-change test. These comparisons are interpreted exclusively as indicators of relative descriptive fit and not as evidence of theoretical superiority, temporal precedence, or causal influence.

Robustness was evaluated through five sensitivity analyses: (1) exclusion of influential observations identified using Cook’s distance (>4/n), (2) alternative heteroscedasticity-consistent standard error estimators (HC1–HC4), (3) alternative coding schemes for professional categories, (4) non-parametric alternatives (Spearman correlation and Kruskal–Wallis tests), and (5) alternative diagnostic-gap formulations. Findings remained directionally consistent across specifications.

One participant had missing gender information (0.3%). This observation was excluded only from gender-stratified analyses and retained in all primary analyses. Complete-case analysis was used for regression models.

All analyses were performed using Python 3.12 with SciPy, NumPy, Pandas, and Statsmodels. Statistical significance was evaluated using two-tailed tests with α = 0.05. Consistent with methodological recommendations, post-hoc observed power was not used for inferential interpretation because it is mathematically redundant with observed effect sizes and significance tests (Hoenig & Heisey, 2001). Instead, emphasis was placed on effect sizes and confidence intervals rather than pots-hoc power.

The study is reported in accordance with the STROBE Statement for cross-sectional studies, and the completed checklist is provided as supplementary material.

## 3. Results

### 3.1 Sample Characteristics

The final analytical sample comprised 303 respondents drawn from the pharmaceutical sector. Practicing pharmacists represented the largest subgroup (n = 213, 70.3%), followed by pharmacy students (n = 61, 20.1%) and academics or graduate students (n = 29, 9.6%). Women constituted 72.6% of participants (n = 220), and the majority of respondents were between 20 and 35 years of age (81.8%).

Professional experience varied across the sample, although nearly half of participants reported between one and five years of experience (46.9%). Participant recruitment, eligibility verification, and analytical inclusion are summarized in Supplementary Figure S1.

The sample included practicing pharmacists, pharmacy students, and academics, providing representation from multiple professional groups within the pharmaceutical sector.

### 3.2 Domain Profile: High Attitudes, Low Readiness, Low Practice

Descriptive statistics are presented in Table 1 and visualized in Figure 2.

**Figure 2.**
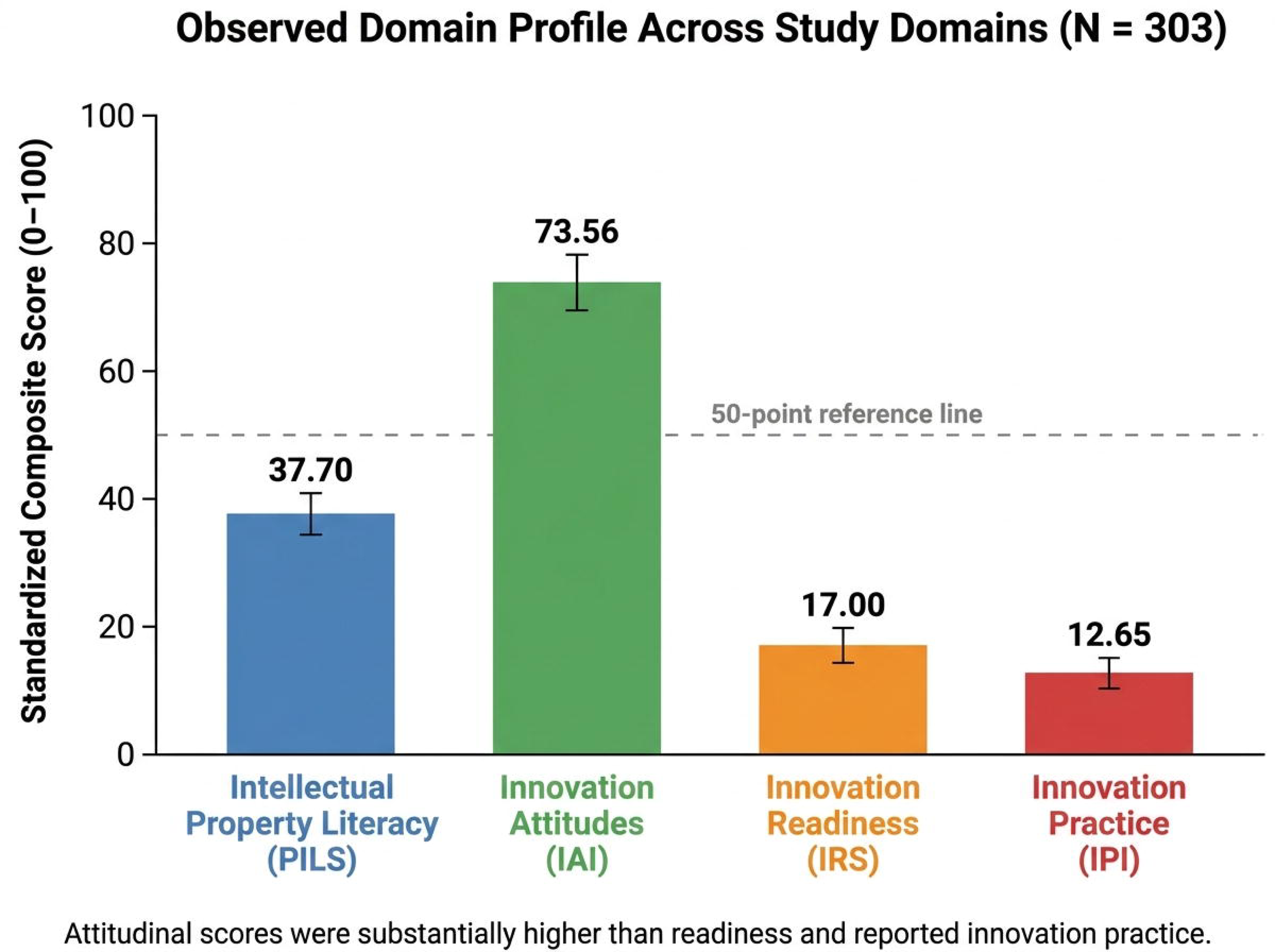
Domain Profile across Study Domains.

**Table 1.** Descriptive Statistics for Study Domains (N = 303)

| Domain | Items | Mean (%) | SD (%) | Range | Reliability |
| --- | --- | --- | --- | --- | --- |
| PILS | 9 | 37.70 | 19.63 | 0–100 | KR-20 = 0.485 |
| IAI | 10 | 73.56 | 6.91 | 54–94 | $\alpha$ = 0.481 |
| IRS | 4 | 17.00 | 22.00 | 0–100 | $\alpha$ = 0.507 |
| IPI | 3 | 12.65 | 22.80 | 0-100 | $\alpha = 0.524$ |
| Diagnostic Gap | — | 60.91 | 23.20 | -40 to 94 | — |

A pronounced imbalance emerged across the four domains. While respondents expressed strong attitudinal support for intellectual property and innovation (IAI = 73.56/100), both innovation readiness (IRS = 17.00/100) and reported innovation practice (IPI = 12.65/100) remained markedly limited. Intellectual property literacy occupied an intermediate position (PILS = 37.70/100).

The resulting arithmetic difference between attitudes and reported practice averaged 60.91 points, indicating a substantial separation between attitudinal endorsement and behavioral expression within the observed system.

The largest contrast was observed between innovation attitudes and reported innovation practice, with a mean difference of 60.91 points. Descriptive statistics are presented in Table 1 and visualized in Figure 2.

### 3.3 Inter-Domain Correlation Structure

The correlation matrix is presented in Table 2 and summarized visually in Figure 3. Knowledge and attitudes were not significantly correlated (r = 0.066, p = 0.253).

**Figure 3.**
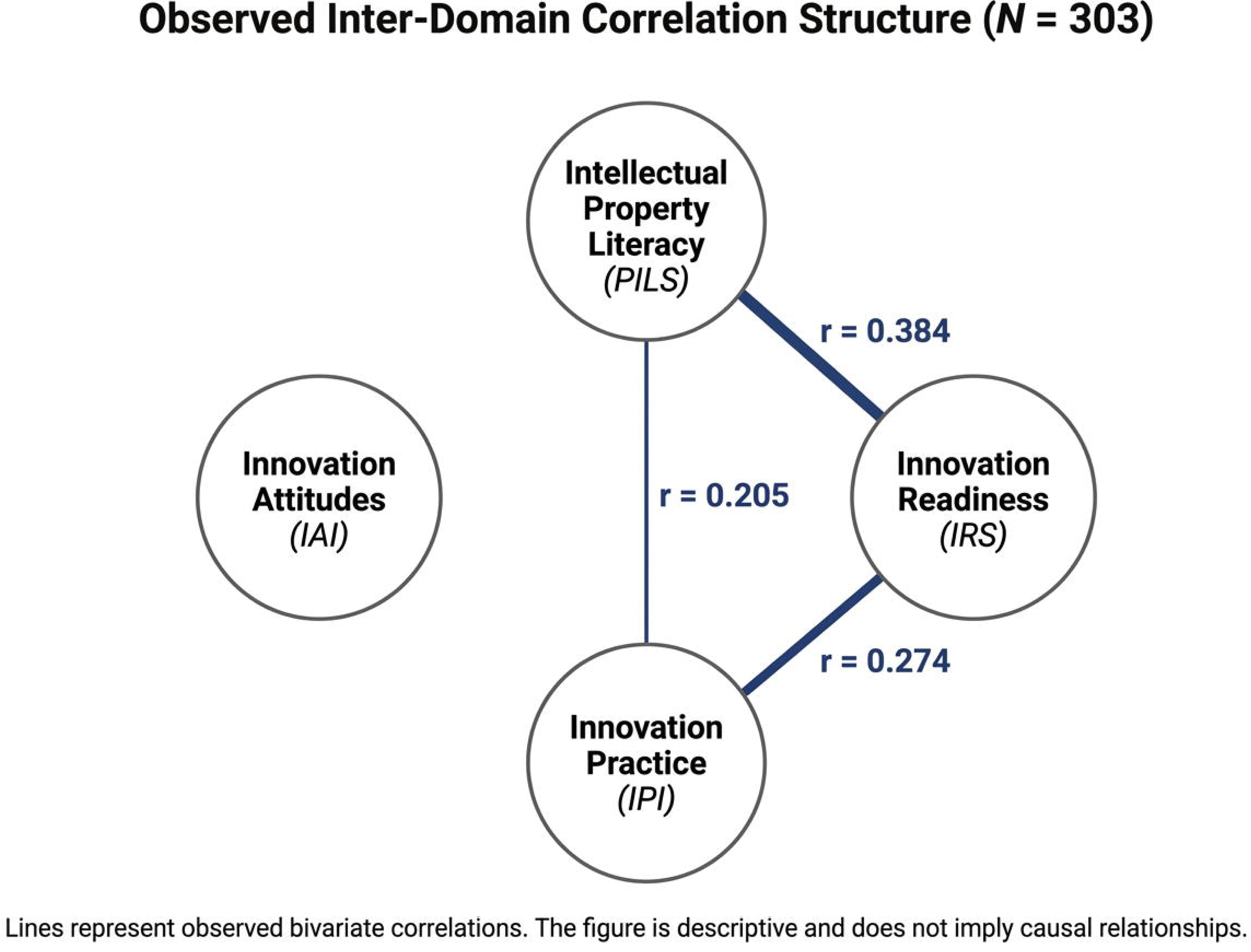
Inter-Domain Correlation Structure (N= 303).

**Table 2.** Inter-Domain Correlation Matrix (N = 303)

| Variable | PILS | IAI | IRS | IPI |
| --- | --- | --- | --- | --- |
| PILS | — | 0.066 (0.253) | 0.384***<br>($<0.001$) | 0.205***<br>($<0.001$) |
| IAI | 0.066 (0.253) | — | 0.104 (0.070) | 0.093 (0.106) |
| IRS | 0.384***<br>($<0.001$) | 0.104 (0.070) | — | 0.274***<br>($<0.001$) |
| IPI | 0.205***<br>($<0.001$) | 0.093 (0.106) | 0.274***<br>($<0.001$) | — |

*Note.* Pearson $r$ reported; $p$ -values in parentheses. \*\*\* $p < 0.001$ .

In contrast, intellectual property literacy demonstrated statistically significant associations with both innovation readiness (r = 0.384, 95% CI [0.27, 0.49], p < 0.001) and innovation practice (r = 0.205, 95% CI [0.09, 0.31], p < 0.001). The strongest observed association involved readiness and practice (r = 0.274, 95% CI [0.16, 0.38], p < 0.001).

Attitudes showed no statistically detectable association with practice (r = 0.093, p = 0.106) and only weak association with readiness (r = 0.104, p = 0.070).

Overall, IP literacy, innovation readiness, and innovation practice showed significant positive associations, whereas innovation attitudes were not significantly associated with reported innovation practice.

### 3.4 Professional Category Comparisons

Table 3 summarizes domain comparisons across pharmacists, students, and academics.

**Table 3.**
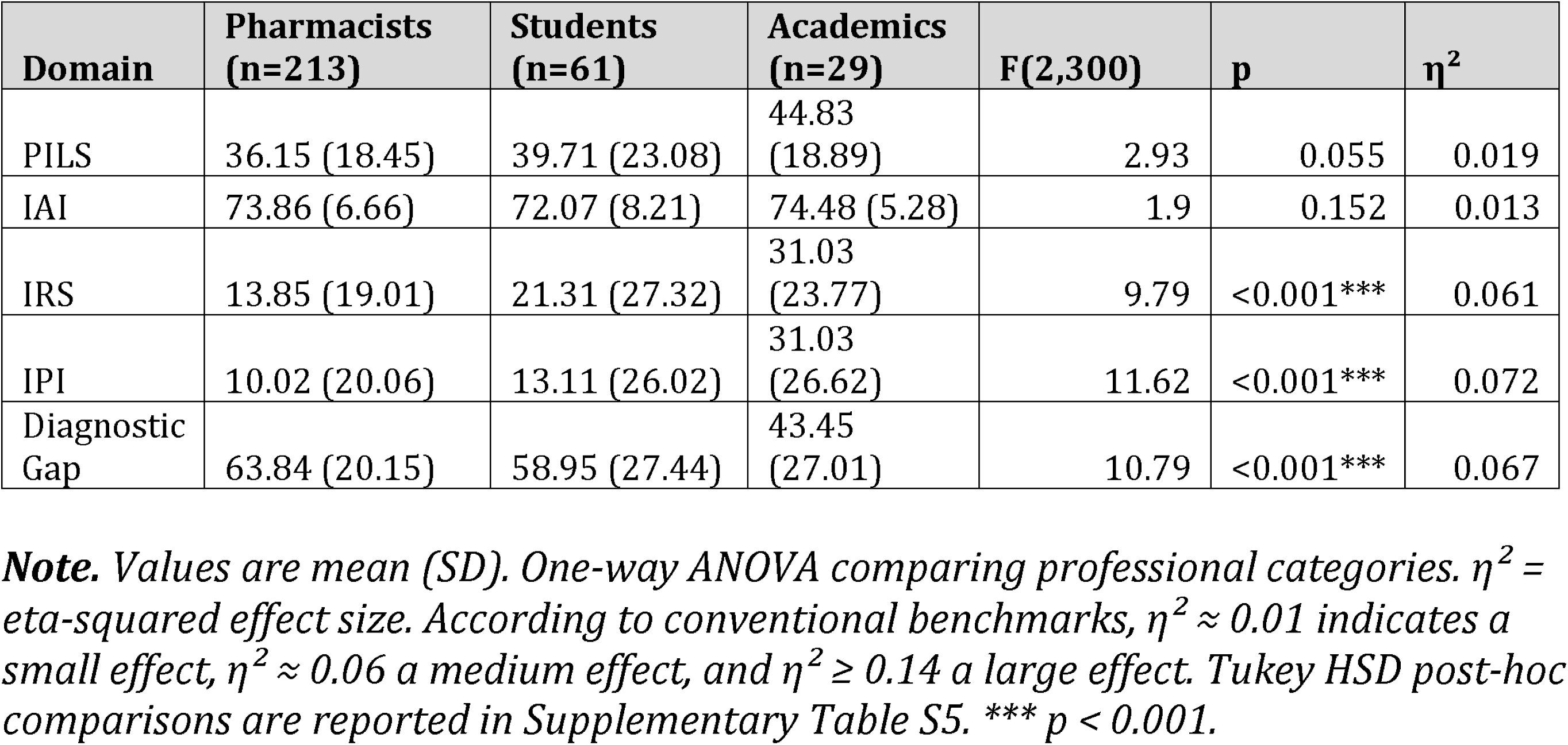
Domain Comparisons across Professional Categories (N = 303)

| Domain | Pharmacists<br>(n=213) | Students<br>(n=61) | Academics<br>(n=29) | F(2,300) | p | $\eta^2$ |
| --- | --- | --- | --- | --- | --- | --- |
| PILS | 36.15 (18.45) | 39.71 (23.08) | 44.83<br>(18.89) | 2.93 | 0.055 | 0.019 |
| IAI | 73.86 (6.66) | 72.07 (8.21) | 74.48 (5.28) | 1.9 | 0.152 | 0.013 |
| IRS | 13.85 (19.01) | 21.31 (27.32) | 31.03<br>(23.77) | 9.79 | <0.001*** | 0.061 |
| IPI | 10.02 (20.06) | 13.11 (26.02) | 31.03<br>(26.62) | 11.62 | <0.001*** | 0.072 |
| Diagnostic<br>Gap | 63.84 (20.15) | 58.95 (27.44) | 43.45<br>(27.01) | 10.79 | <0.001*** | 0.067 |

**Note.** Values are mean (SD). One-way ANOVA comparing professional categories. $\eta^2 =$ eta-squared effect size. According to conventional benchmarks, $\eta^2 \approx 0.01$ indicates a small effect, $\eta^2 \approx 0.06$ a medium effect, and $\eta^2 \geq 0.14$ a large effect. Tukey HSD post-hoc comparisons are reported in Supplementary Table S5. \*\*\* $p < 0.001$ .

No statistically significant differences were observed for intellectual property literacy (F = 2.93, p = 0.055) or innovation attitudes (F = 1.90, p = 0.152), indicating broadly similar knowledge and attitudinal profiles across professional categories.

In contrast, significant differences were observed for innovation readiness (F = 9.79, p < 0.001), innovation practice (F = 11.62, p < 0.001), and the diagnostic gap indicator (F = 10.79, p < 0.001). Academics achieved the highest readiness and innovation practice scores, whereas practicing pharmacists exhibited the largest attitude–practice gap.

Post-hoc comparisons (Supplementary Table S5) showed that the largest differences involved academics, who scored significantly higher than practicing pharmacists on both readiness and innovation practice measures.

Effect-size estimates indicated small effects for intellectual property literacy (η² = 0.019) and innovation attitudes (η² = 0.013), whereas moderate effects were observed for innovation readiness (η² = 0.061), innovation practice (η² = 0.072), and the diagnostic gap indicator (η² = 0.067).

### 3.5 Comparative Specification Models

To further characterize the observed domain configuration, two comparative specifications were estimated using HC3-robust regression models (Table 4).

**Table 4.** Comparative Specification Models (HC3 Robust Standard Errors)

| Predictor | Specification A<br>(IRS) $\beta$ (SE) | Spec A p | Specification B<br>(IPI) $\beta$ (SE) | Spec B p |
| --- | --- | --- | --- | --- |
| Constant | -20.61 (15.08) | 0.173 | -15.80 (14.74) | 0.286 |
| PILS | 0.40 (0.07) | <0.001*** | 0.12 (0.08) | 0.140 |
| IAI | 0.30 (0.18) | 0.099 | 0.20 (0.20) | 0.332 |
| IRS | — | — | 0.19 (0.09) | 0.028* |
| Gender (Male) | 3.57 (2.95) | 0.226 | -2.33 (2.99) | 0.436 |
| Student | 5.20 (4.15) | 0.210 | 4.04 (4.58) | 0.378 |
| Academic | 13.98 (3.96) | <0.001*** | 15.53 (5.13) | 0.002** |
| Experience Level | -1.31 (1.40) | 0.349 | 2.10 (1.63) | 0.196 |

*Note. HC3 robust standard errors. \*\*\* $p < 0.001$ ; \*\* $p < 0.01$ ; \* $p < 0.05$ .*

**Table 4b.** Standardized Coefficients and Partial R² for Predictor Importance Comparison.

| Predictor | Specification<br>A (IRS) $\beta^*$ | Spec A<br>partial $R^2$ | Spec A<br>Rank | Specification<br>B (IPI) $\beta^*$ | Spec B<br>partial $R^2$ | Spec B<br>Rank |
| --- | --- | --- | --- | --- | --- | --- |
| PILS | 0.357 | 0.099 | 1 | 0.103 | 0.008 | 3 |
| IAI | 0.094 | 0.009 | 4 | 0.061 | 0.003 | 5 |
| IRS | — | — | — | 0.183 | 0.015 | 2 |
| Gender<br>(Male) | 0.072 | 0.005 | 5 | -0.046 | 0.002 | 6 |
| Student | 0.095 | 0.005 | 3 | 0.071 | 0.003 | 4 |
| Academic | 0.187 | 0.040 | 2 | 0.200 | 0.030 | 1 |
| Experience Level | — | 0.003 | 6 | — | 0.006 | 7 |

*Note. $\beta$ = standardized beta coefficient. Partial $R^2$ = unique variance explained by each predictor after accounting for all other predictors in the specification. Rank = relative importance based on partial $R^2$ . Experience Level partial $R^2$ computed from $t = -0.936$ (Spec A) and $t = 1.288$ (Spec B).*

#### Specification A: Innovation Readiness

The first specification examined variables associated with innovation readiness (IRS). Intellectual property literacy emerged as the strongest predictor of readiness (β = 0.40, 95% CI [0.25, 0.55], p < 0.001), while academic status was also positively associated with higher readiness levels (β = 13.98, 95% CI [6.00, 21.96], p < 0.001).

Together, the included variables accounted for 19.9% of observed variation in readiness (R² = 0.199).

#### Specification B: Innovation Practice

The second specification examined reported innovation practice (IPI) while incorporating readiness as an additional explanatory variable.

Innovation readiness was positively associated with innovation practice (β = 0.19, 95% CI [0.02, 0.36], p = 0.028), as was academic status (β = 15.53, 95% CI [5.50, 25.56], p = 0.002).

Notably, the previously significant bivariate association between intellectual property literacy and innovation practice was attenuated and no longer statistically detectable after readiness was introduced into the same specification (β = 0.12, p = 0.140). Attitudes remained non-significant throughout (β = 0.20, p = 0.332).

After innovation readiness was included in the model, the association between IP literacy and innovation practice was attenuated and no longer statistically significant (β = 0.12, p = 0.140). Innovation attitudes remained non-significant in all model specifications.

### 3.6 Comparative Assessment of Knowledge–Attitude and Readiness-Oriented Specifications

To evaluate the descriptive contribution of readiness, a baseline specification containing knowledge and attitudes was compared with a readiness-expanded specification (Table 5).

**Table 5.** Comparative Specification Assessment: Baseline vs Readiness-Expanded (Predicting IPI)

| Specification | Predictors | $R^2$ | AIC | BIC |
| --- | --- | --- | --- | --- |
| Baseline (K) | PILS + IAI + covariates | 0.082 | 1,247.3 | 1,268.1 |
| Readiness-Expanded (R) | PILS + IAI + IRS + covariates | 0.140 | 1,234.9 | 1,259.4 |
| Difference (R – K) | — | $\Delta R^2 = 0.058^*$ | $\Delta AIC = -12.4$ | $\Delta BIC = -8.7$ |

*Note. $*p < 0.05$ for $\Delta R^2$ . Covariates: Gender, Student, Academic, Experience Level.*

The baseline model explained 8.2% of observed variation in innovation practice (R² = 0.082). Introducing readiness increased explained variation to 14.0% (R² = 0.140), yielding an incremental improvement of ΔR² = 0.058 (p = 0.028).

Model fit also improved according to both information criteria (ΔAIC = −12.4; ΔBIC = −8.7).

Importantly, the inclusion of readiness coincided with attenuation of the literacy–practice association observed in the bivariate analysis. The readiness-expanded specification demonstrated higher explanatory fit than the baseline knowledge–attitude model, as indicated by increases in R² and improvements in AIC and BIC values.

### 3.7 Model Diagnostics and Robustness

Diagnostic evaluation revealed no evidence of problematic multicollinearity, with all variance inflation factors remaining below 2.0.

Residual analyses indicated departures from normality and the presence of heteroscedasticity, both of which are expected when modeling bounded composite scores. Accordingly, HC3 heteroscedasticity-consistent standard errors were retained throughout the primary analyses.

Sensitivity analyses using alternative standard-error estimators, exclusion of influential observations, alternative coding schemes, non-parametric procedures, and alternative gap formulations produced directionally consistent findings (Supplementary Table S7), supporting the robustness of the reported patterns.

### 3.8 Convergent Validity Assessment

To evaluate convergent validity of the intellectual property literacy measure, respondents who self-identified as knowledgeable about intellectual property (Q10 = Yes) were compared with those who did not.

Participants reporting perceived knowledge achieved significantly higher objective literacy scores than those reporting limited knowledge (4.00 ± 1.90 vs. 2.75 ± 1.34; p < 0.001), corresponding to a moderate point-biserial association (rpb = 0.355).

Respondents reporting prior knowledge of intellectual property achieved significantly higher objective literacy scores, providing evidence of convergent validity for the PILS measure.

## 4. Discussion

### 4.1 Beyond Knowledge and Attitudes: Understanding Innovation under Constraint

The present study set out to examine how intellectual property literacy, innovation attitudes, innovation readiness, and reported innovation practice co-occur within a transitional pharmaceutical system. The resulting empirical configuration reveals a pattern that differs substantially from conventional knowledge-centered interpretations of innovation behavior.

The most notable finding is the coexistence of relatively strong attitudinal endorsement of innovation with very limited readiness and reported innovation practice. While participants generally expressed favorable views toward intellectual property and innovation-related activities, these orientations were not accompanied by corresponding levels of behavioral engagement. This divergence suggests that positive attitudes alone provide only limited information about whether innovation-related activities are practically feasible within constrained institutional environments.

At the same time, knowledge was positively associated with both readiness and practice, indicating that cognitive resources remain relevant components of innovation systems. Yet knowledge alone did not appear sufficient to account for observed behavioral variation. Instead, the pattern suggests that the translation of knowledge into practice may depend on the presence of enabling conditions that make innovation-related activity operationally possible. Similar gaps between awareness, attitudes, and implementation have been reported in pharmaceutical and healthcare contexts, where favorable perceptions often coexist with limited institutional capacity for execution (Odeh et al., 2022; Wang et al., 2023).

Viewed collectively, the findings point toward a system characterized not by a lack of awareness or attitudinal support, but by limited capacity for behavioral translation.

### 4.2 Innovation Readiness as a Proxy for Enabling Conditions

A central finding of this study is the role of innovation readiness as an indicator of contextual conditions associated with innovation-related activity. Readiness was operationalized using observable indicators reflecting access to innovation infrastructure, exposure to research environments, organizational support, and innovation-related training opportunities.

Across analyses, readiness showed a stronger relationship with reported innovation practice than did innovation attitudes. Moreover, the inclusion of readiness improved model fit beyond knowledge–attitude specifications alone. In the multivariable analyses, readiness remained significantly associated with innovation practice, whereas attitudes did not demonstrate a statistically significant relationship.

These findings should not be interpreted as evidence of causality, mediation, or temporal ordering. Nevertheless, they suggest that contextual conditions may provide information about innovation-related behavior that is not captured by knowledge or attitudinal measures alone. This interpretation is consistent with innovation-systems research emphasizing the importance of institutional and organizational environments in shaping the practical application of knowledge resources (OECD, 2018; Van den Hoed et al., 2022).

Taken together, the findings indicate that innovation-related knowledge and favorable attitudes may coexist with limited innovation activity when opportunities for engagement, collaboration, and implementation remain constrained.

### 4.3 A Relationships among Knowledge, Attitudes, Readiness, and Practice

The observed pattern of associations differed from the assumptions commonly implied by linear knowledge–attitude–practice frameworks. Knowledge was positively associated with both readiness and innovation practice, whereas attitudes were not significantly associated with practice. In addition, readiness demonstrated a positive association with innovation practice and contributed additional explanatory information in the comparative specification analyses.

These results suggest that the four domains examined in this study are not equally related to innovation-related behavior. While knowledge appears to remain relevant, readiness showed a closer empirical relationship with reported practice than attitudinal measures. This finding highlights the potential importance of contextual conditions when interpreting innovation-related outcomes in resource-constrained environments.

At the same time, a substantial proportion of variation in innovation practice remained unexplained. This observation indicates that additional factors, including organizational culture, regulatory conditions, resource availability, collaboration networks, and broader institutional influences, may also contribute to innovation-related behavior and warrant further investigation in future studies.

### 4.4 Implications for Pharmaceutical Innovation Policy

The policy relevance of these findings lies in the distinction between awareness and implementation. Efforts aimed exclusively at improving knowledge or strengthening positive attitudes may not be sufficient when the underlying conditions required for innovation remain weak.

Within the present sample, respondents already exhibited relatively favorable attitudes toward innovation and intellectual property. Nevertheless, readiness and practice scores remained low. This pattern suggests that interventions focused solely on educational dissemination may encounter diminishing returns if institutional bottlenecks continue to limit opportunities for practical engagement.

Instead, the findings indicate that capability-oriented strategies may provide a more informative diagnostic lens for understanding innovation performance in constrained environments. Such strategies could include strengthening innovation infrastructure, expanding exposure to research ecosystems, increasing organizational support mechanisms, and creating institutional pathways that facilitate the practical application of knowledge.

The present findings may be particularly relevant for Syria, where a substantial pharmaceutical manufacturing base coexists with a large pharmacy education sector. Although the current study was not designed to evaluate educational interventions, the observed gap between innovation-related knowledge and innovation practice suggests that greater emphasis on innovation competencies, intellectual property management, entrepreneurship, and research translation may warrant consideration. Such an approach aligns with international innovation-capacity frameworks promoted by the World Intellectual Property Organization (WIPO), which emphasize human-capital development, innovation skills, and intellectual property education as important components of innovation ecosystems (WIPO Academy 2026; WIPO Global Innovation Index 2025). Future research should examine whether integrating these elements into pharmaceutical education and professional development programs contributes to measurable improvements in innovation readiness and innovation practice These observations should be interpreted cautiously. The study does not demonstrate that readiness causes innovation practice, nor does it establish any temporal ordering among domains. Rather, the results highlight a recurring empirical pattern in which capability-related conditions appear more closely aligned with reported innovation activity than attitudinal measures alone. From a policy standpoint, this suggests that strengthening enabling environments may deserve equal attention alongside knowledge-development initiatives when seeking to improve innovation outcomes in transitional pharmaceutical systems.

### 4.5 Study Strengths

Several strengths enhance the contribution of the present study. First, the study introduces innovation readiness as an empirically observable capability-oriented domain within pharmaceutical innovation research. Second, the comparative specification approach allows evaluation of alternative descriptive representations of innovation-related behavior without imposing causal assumptions. Third, the inclusion of professionals, students, and academics provides a heterogeneous perspective on innovation-related conditions across different stages of professional engagement. Finally, extensive robustness and sensitivity analyses support the stability of the observed empirical patterns.

### 4.6 Limitations

Several limitations should be acknowledged. First, the cross-sectional design precludes assessment of temporal ordering among domains. Second, convenience sampling may limit external generalizability. Third, all measures relied on self-reported responses and may therefore be influenced by reporting bias. Fourth, the readiness and practice indicators were intentionally operationalized as composite descriptive measures rather than validated latent constructs. Finally, the study was conducted within a single national context, and replication across multiple institutional settings is required before broader generalizations can be made.

## 5. Conclusions

This study examined the relationships among intellectual property literacy, innovation attitudes, innovation readiness, and reported innovation practice within Syria’s pharmaceutical sector. The findings revealed a consistent pattern in which innovation attitudes were comparatively high, whereas innovation readiness and reported innovation practice remained substantially lower.

Although intellectual property literacy was positively associated with both readiness and practice, innovation attitudes showed little relationship with reported innovation activity. In contrast, innovation readiness demonstrated a closer association with practice and provided additional descriptive information beyond conventional knowledge–attitude approaches. These findings suggest that innovation-related behavior may be more closely linked to enabling conditions than to favorable attitudes alone.

At the same time, a substantial proportion of variation in innovation practice remained unexplained, indicating that innovation activity is influenced by a broader set of organizational, institutional, and contextual factors that were not captured in the present study.

Given the cross-sectional design, all findings should be interpreted as descriptive associations rather than evidence of causality, mediation, or temporal ordering. Future research should examine these relationships across different institutional settings and through longitudinal designs to determine whether the observed patterns remain stable over time.

Overall, the study highlights the importance of considering contextual enabling conditions alongside knowledge and attitudes when assessing innovation capacity in resource-constrained pharmaceutical systems. The findings suggest that strengthening opportunities, support structures, and innovation-related environments may be as important as improving knowledge and awareness when seeking to translate innovation potential into observable practice.

From a practical perspective, the observed gap between innovation-related capabilities and actual innovation practice highlights the potential value of strengthening innovation-oriented education and innovation-supportive ecosystems within pharmaceutical training and professional development. In Syria, where a large network of pharmacy faculties coexists with an established pharmaceutical manufacturing sector, strengthening education in innovation, intellectual property, entrepreneurship, and pharmaceutical product development may help cultivate systematic innovation competencies among future pharmacists and pharmaceutical professionals. Such efforts could facilitate the translation of academic research and experimental work into practical applications, promote stronger university–industry engagement, and enhance opportunities for innovation-related participation.

The broader context of the present study also highlights growing interest in intellectual property awareness and innovation-related engagement within the pharmaceutical community. Data collection coincided with World Intellectual Property Day 2026 outreach activities organized by pharmacy students and academic staff at Manara University, which involved awareness-raising initiatives and discussions related to intellectual property and pharmaceutical innovation. Although these activities were not formally evaluated within the study framework, they illustrate the potential role of educational and awareness-oriented initiatives in fostering dialogue around innovation, intellectual property, and research translation.

While the effectiveness of such educational and institutional interventions was not examined in the present study and requires future evaluation, they represent plausible strategies for improving innovation readiness and strengthening the conditions that support innovation practice within Syria’s pharmaceutical sector.

## Data Availability

All relevant data underlying the findings of this study are publicly available. The de-identified dataset, complete analysis code, supplementary materials, and reproducibility documentation are available in the GitHub repository and permanently archived on Zenodo (DOI: 10.5281/zenodo.20716882). These materials enable full reproduction of all analyses reported in the manuscript.

https://doi.org/10.5281/zenodo.20716882

https://github.com/chadikhatib/innovation-capability-constraints

## Supplementary Materials

### - Supplementary Figure

**Figure S1.**
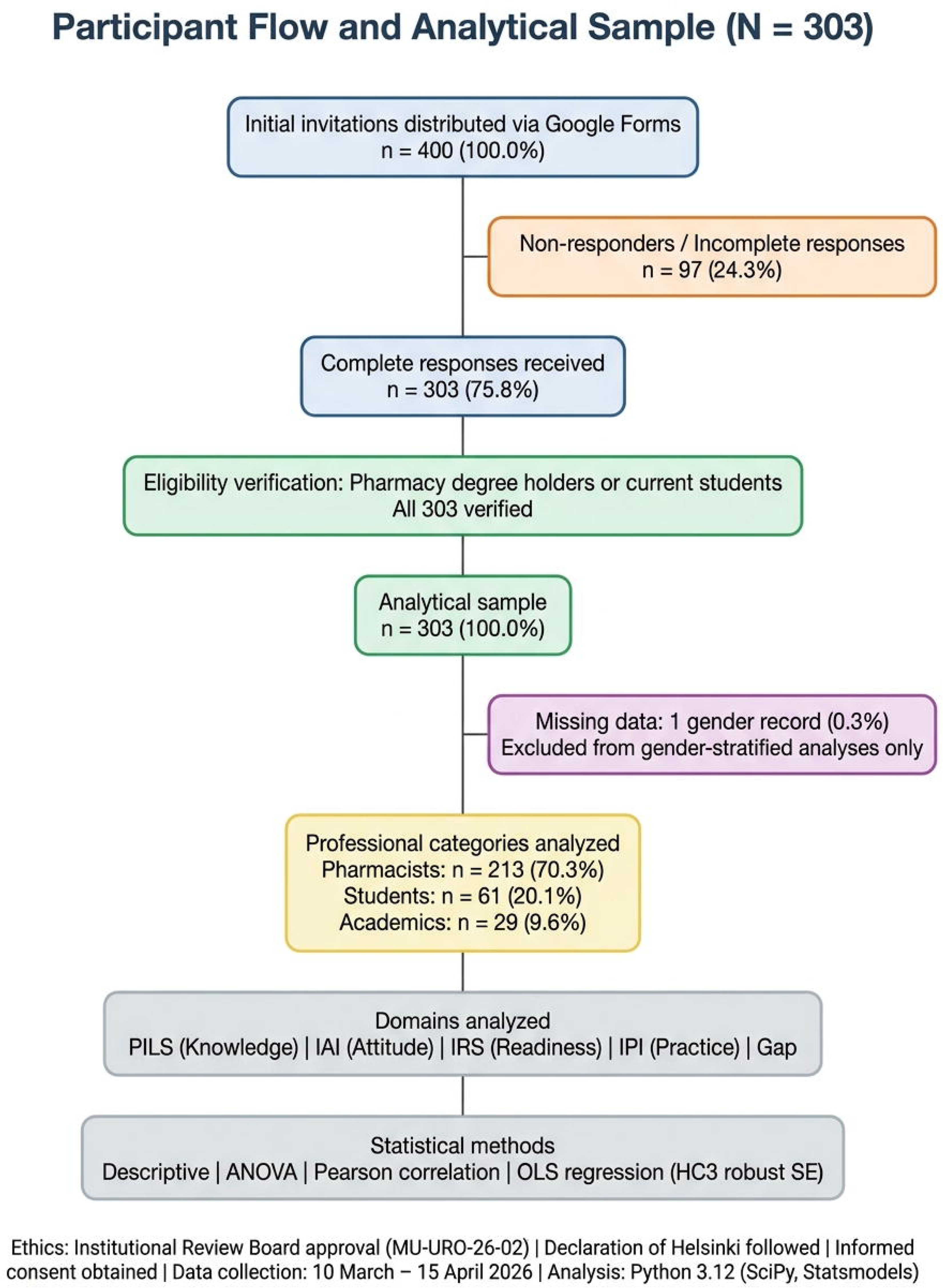
Participant Flow Diagram.

### - Supplementary Tables

**Table S1.** Participant Flow and Analytical Sample Summary

**Table S2.** Demographic and Professional Characteristics (N = 303)

**Table S3.** Item-Level Descriptive Statistics

**Table S4.** Inter-Domain Pearson Correlations with 95% Confidence Intervals

**Table S5.** ANOVA Post-Hoc Comparisons (Tukey HSD)

**Table S6.** Regression Diagnostics and Nested Model Comparison

**Table S7.** Sensitivity Analysis Results

**Table S8.** Convergent Validity: Self-Assessment (Q10) vs PILS Availability of Data and Materials

The de-identified dataset (N = 303), complete analysis code, supplementary tables (S1–S8), statistical analysis plan (SAP v1.0), STROBE checklist, and reproducibility materials are publicly available in <u>GitHub repository</u> the and permanently archived in <u>Zenodo</u> (doi:10.5281/zenodo.20716882). The dataset is provided in CSV format with synthetic participant identifiers (P001–P303) and contains no personally identifiable information. All analyses are fully reproducible using Python 3.12+ with dependencies specified in requirements.txt. A citation file (CITATION.cff) is included to facilitate proper attribution (Khatib et al., 2026)

## AI Disclosure

The authors used AI-assisted writing tools (large language models) for language refinement and structural organization. All scientific content, data interpretation, statistical analysis, clinical conclusions, and ethical oversight were authored and verified by the research team. No AI tool was used for data analysis, statistical computation, or generation of original research findings.

## Ethics Approval and Consent to Participate

Ethical approval was obtained from the institutional review board of the authors’ institution (Approval No. MU-URO-26-02). The study was conducted in accordance with the Declaration of Helsinki.

Written informed consent was obtained electronically from all participants prior to participation via a mandatory consent checkbox before accessing the questionnaire.

## Author Contributions

**CK** conceptualized and supervised the study. **CK** contributed to methodology, formal analysis, and writing – review & editing. **HA, ZH, MI**, and **JM** contributed to investigation, data curation, formal analysis, and writing – original draft. All authors read and approved the final manuscript.

## Acknowledgments

The authors gratefully acknowledge the institutional support and commitment to research excellence provided by the President of Manara University, the University Administration, and the Dean of the Faculty of Pharmacy.

The authors extend their sincere appreciation to the Syrian Pharmacists Syndicate, the Syrian Medical Syndicate, Latakia Hospital, Latakia University, and Al-Sham University for their logistical and professional support during the conduct of this study.

The authors are particularly grateful to the Syrian Association for Intellectual Property and Development (Mr. Yasser Saada) for their advisory contributions, and to Al-Manara Law Firm, Consultancies and Legal Studies, and specifically to General Manager and Legal Counsel Mr. Ahmad Talal Anis Karknawi for his expert legal guidance.

Finally, the authors wish to express their profound gratitude to all study volunteers and survey participants who generously devoted their time to complete the questionnaire on a voluntary basis. This research would not have been possible without their willingness to contribute.

The authors acknowledge the pharmacy students and academic staff of Manara University who participated in World Intellectual Property Day 2026 awareness activities conducted during the study period.

