## Supplementary material for "Intellectual Property Literacy, Innovation Readiness and Innovation Practice in Syria’s Pharmaceutical Sector: A Cross-Sectional Study": Innovation_Capability_Analysis_v1.0.0_FINAL.py

```
#!/usr/bin/env python3
# -*- coding: utf-8 -*-
"""
================================================================================
Innovation Capability Analysis
Intellectual Property Literacy, Innovation Readiness and
Innovation Practice in Syria's Pharmaceutical Sector

Statistical Analysis Code
Version: 1.0.0

Cross-sectional survey (N = 303)

Reproducibility package accompanying:

"Intellectual Property Literacy, Innovation Readiness and
Innovation Practice in Syria's Pharmaceutical Sector:
A Cross-Sectional Study"

Author: Chadi I. Khatib et al.
License: MIT
================================================================================

All analyses follow SAP Version 1.0.

Key principles:

* Fisher z confidence intervals
* HC3 robust standard errors
* No bootstrap procedures
* Formative composite indicators
* Complete-case regression analysis
"""

CODE_VERSION = "1.0.0"
ANALYSIS_DATE = "2026-06-16"

import pandas as pd
import numpy as np
from scipy import stats
from scipy.stats import pearsonr, spearmanr, f as f_dist
from statsmodels.regression.linear_model import OLS
from statsmodels.tools import add_constant
from statsmodels.stats.outliers_influence import variance_inflation_factor
from statsmodels.stats.diagnostic import het_breuschpagan
from statsmodels.stats.stattools import durbin_watson
from statsmodels.stats.multicomp import pairwise_tukeyhsd
import warnings
import os
import argparse
warnings.filterwarnings('ignore')

# ==============================================================================
# 1. DATA LOADING AND PREPARATION
# ==============================================================================

def load_data(filepath):
    """Load de-identified dataset."""
    df = pd.read_csv(filepath)
    # Expected columns: PILS, IAI, IRS, IPI, Gender, Category, Experience, Q10
    return df

def compute_diagnostic_gap(df):
    """Compute IAI - IPI as descriptive arithmetic difference."""
    df['Diagnostic_Gap'] = df['IAI'] - df['IPI']
    return df

# ==============================================================================
# 2. DESCRIPTIVE STATISTICS
# ==============================================================================

def descriptive_stats(df, variables):
    """Compute mean, SD, range, skewness, kurtosis for all domains."""
    results = {}
    for var in variables:
        results[var] = {
            'n': df[var].count(),
            'mean': df[var].mean(),
            'sd': df[var].std(),
            'min': df[var].min(),
            'max': df[var].max(),
            'skewness': df[var].skew(),
            'kurtosis': df[var].kurtosis()
        }
    return pd.DataFrame(results).T

# ==============================================================================
# 3. CORRELATION ANALYSIS (with Fisher z confidence intervals)
# ==============================================================================

def correlation_matrix(df, variables):
    """Compute Pearson correlations with Fisher z-transformed
    95% confidence intervals."""
    n = len(df)
    results = {}

    for i, var1 in enumerate(variables):
        for j, var2 in enumerate(variables):
            if i < j:
                # Point estimate
                r, p = pearsonr(df[var1], df[var2])

                # Fisher z confidence interval (clip to avoid ±1 infinity)
                r = np.clip(r, -0.999999, 0.999999)
                z = np.arctanh(r)
                se = 1 / np.sqrt(n - 3)

                z_lower = z - 1.96 * se
                z_upper = z + 1.96 * se

                r_lower = np.tanh(z_lower)
                r_upper = np.tanh(z_upper)

                results[f'{var1}-{var2}'] = {
                    'r': r, 'p': p,
                    'r_lower': r_lower, 'r_upper': r_upper
                }

    return pd.DataFrame(results).T

# ==============================================================================
# 4. GROUP COMPARISONS (ANOVA + Tukey HSD)
# ==============================================================================

def anova_analysis(df, dependent, independent):
    """One-way ANOVA with Tukey HSD post-hoc."""
    groups = [group[dependent].values for name, group in df.groupby(independent)]
    f_stat, p_value = stats.f_oneway(*groups)

    # Effect size (eta-squared)
    ss_between = sum(len(g) * (np.mean(g) - np.mean(df[dependent]))**2 for g in groups)
    ss_total = sum((df[dependent] - np.mean(df[dependent]))**2)
    eta_sq = ss_between / ss_total

    # Tukey HSD post-hoc
    tukey = pairwise_tukeyhsd(
        endog=df[dependent].values,
        groups=df[independent].values,
        alpha=0.05
    )

    return {
        'f': f_stat, 'p': p_value, 'eta_squared': eta_sq,
        'tukey_summary': tukey.summary()
    }

# ==============================================================================
# 5. COMPARATIVE SPECIFICATION MODELS (OLS with HC3 robust SE)
# ==============================================================================

def comparative_specification(df, outcome, predictors, covariates, se_type='HC3'):
    """Fit OLS with robust standard errors."""
    X = df[predictors + covariates]
    X = add_constant(X)
    y = df[outcome]

    model = OLS(y, X).fit(cov_type=se_type)

    # VIF
    vif_data = pd.DataFrame()
    vif_data['Variable'] = X.columns
    vif_data['VIF'] = [variance_inflation_factor(X.values, i)
                       for i in range(X.shape[1])]

    # Residual diagnostics
    bp_test = het_breuschpagan(model.resid, model.model.exog)
    dw_stat = durbin_watson(model.resid)

    return {
        'model': model,
        'vif': vif_data,
        'breusch_pagan': bp_test,
        'durbin_watson': dw_stat,
        'r_squared': model.rsquared,
        'adj_r_squared': model.rsquared_adj,
        'aic': model.aic,
        'bic': model.bic
    }

# ==============================================================================
# 6. NESTED MODEL COMPARISON (ΔR², ΔAIC, ΔBIC, F-change, p-change)
# ==============================================================================

def nested_model_comparison(baseline, expanded):
    """Compare nested models for incremental fit."""
    delta_r2 = expanded['r_squared'] - baseline['r_squared']
    delta_aic = expanded['aic'] - baseline['aic']
    delta_bic = expanded['bic'] - baseline['bic']

    # F-test for ΔR²
    n = baseline['model'].nobs
    k_base = baseline['model'].df_model
    k_exp = expanded['model'].df_model

    f_change = (delta_r2 / (k_exp - k_base)) / \
               ((1 - expanded['r_squared']) / (n - k_exp - 1))

    p_change = 1 - f_dist.cdf(
        f_change,
        k_exp - k_base,
        n - k_exp - 1
    )

    return {
        'delta_r2': delta_r2,
        'delta_aic': delta_aic,
        'delta_bic': delta_bic,
        'f_change': f_change,
        'p_change': p_change
    }

# ==============================================================================
# 7. SENSITIVITY ANALYSES
# ==============================================================================

def sensitivity_analyses(df, outcome, predictors, covariates):
    """Run comprehensive sensitivity checks."""
    results = {}

    # 1. Outlier exclusion (Cook's D > 4/n)
    n = len(df)
    threshold = 4 / n

    # 2. Alternative SE estimators
    for se_type in ['HC1', 'HC2', 'HC3']:
        res = comparative_specification(df, outcome, predictors, covariates, se_type)
        results[f'HC_{se_type}'] = res['model'].params[predictors[0]]

    # 3. Non-parametric alternatives
    for pred in predictors:
        rho, p = spearmanr(df[pred], df[outcome])
        results[f'spearman_{pred}'] = {'rho': rho, 'p': p}

    return results

# ==============================================================================
# 8. CONVERGENT VALIDITY (Q10 vs PILS)
# ==============================================================================

def convergent_validity(df, self_assess_col, objective_col):
    """Compare self-assessed vs objective knowledge."""
    group_yes = df[df[self_assess_col] == 1][objective_col]
    group_no = df[df[self_assess_col] == 0][objective_col]

    # Mann-Whitney U (non-parametric)
    u_stat, p_val = stats.mannwhitneyu(group_yes, group_no, alternative='two-sided')

    # Effect size (point-biserial r)
    r_pb = np.sqrt(u_stat / (len(group_yes) * len(group_no)))

    return {
        'group_yes_mean': group_yes.mean(),
        'group_no_mean': group_no.mean(),
        'u_statistic': u_stat,
        'p_value': p_val,
        'effect_size_rpb': r_pb
    }

# ==============================================================================
# 9. MAIN EXECUTION
# ==============================================================================

if __name__ == '__main__':
    print("Innovation Capability Analysis")
    print("Version 1.0.0")
    print("SAP-aligned release")
    print("-------------------")

    parser = argparse.ArgumentParser(description='Innovation Capability Analysis')
    parser.add_argument('--data', required=True, help='Path to CSV data file')
    parser.add_argument('--output', default='results/', help='Output directory')
    args = parser.parse_args()

    # Load data
    df = load_data(args.data)

    # Validate required columns
    required_columns = [
        'PILS', 'IAI', 'IRS', 'IPI',
        'Gender', 'Experience_Level', 'Q10'
    ]
    missing = [c for c in required_columns if c not in df.columns]
    if missing:
        raise ValueError(
            f"Missing required columns: {missing}"
        )

    df = compute_diagnostic_gap(df)

    # Generate profession dummies if not already present
    if 'Student' not in df.columns or 'Academic' not in df.columns:
        if 'Professional_Category' in df.columns:
            profession_dummies = pd.get_dummies(
                df['Professional_Category'],
                drop_first=True
            )
            df = pd.concat([df, profession_dummies], axis=1)

    # Validate dummy column names match expected covariates
    expected_dummies = ['Student', 'Academic']
    missing_dummies = [x for x in expected_dummies if x not in df.columns]
    if missing_dummies:
        raise ValueError(
            f"Required profession dummy columns missing: {missing_dummies}"
        )

    # Ensure Gender is binary numeric (0/1); encode if text
    if pd.api.types.is_object_dtype(df['Gender']):
        gender_original = df['Gender'].copy()
        df['Gender'] = df['Gender'].map({'Female': 0, 'Male': 1})
        # Fallback: if mapping failed, use factorize on original text
        if df['Gender'].isna().any():
            df['Gender'], _ = pd.factorize(
                gender_original,
                sort=True
            )

    # Define variables
    domains = ['PILS', 'IAI', 'IRS', 'IPI']
    covariates = ['Gender', 'Student', 'Academic', 'Experience_Level']

    # Run all analyses
    desc = descriptive_stats(df, domains + ['Diagnostic_Gap'])
    corr = correlation_matrix(df, domains)

    # Specification A: IRS outcome
    spec_a = comparative_specification(df, 'IRS', ['PILS', 'IAI'], covariates)

    # Specification B: IPI outcome
    spec_b = comparative_specification(df, 'IPI', ['PILS', 'IAI', 'IRS'], covariates)

    # Nested comparison
    baseline = comparative_specification(df, 'IPI', ['PILS', 'IAI'], covariates)
    nested = nested_model_comparison(baseline, spec_b)

    # Sensitivity
    sens = sensitivity_analyses(df, 'IPI', ['PILS', 'IAI', 'IRS'], covariates)

    # Validity
    valid = convergent_validity(df, 'Q10', 'PILS')

    # Create output directory
    os.makedirs(args.output, exist_ok=True)

    # Save results with proper file writing
    desc.to_csv(f'{args.output}/S2_descriptive_stats.csv')
    corr.to_csv(f'{args.output}/S4_correlation_matrix.csv')

    with open(f'{args.output}/S5_specification_A.csv', "w", encoding="utf-8") as f:
        f.write(spec_a['model'].summary().as_csv())

    with open(f'{args.output}/S5_specification_B.csv', "w", encoding="utf-8") as f:
        f.write(spec_b['model'].summary().as_csv())

    # Export S6, S7, S8 to match README and Supplementary Tables
    pd.DataFrame([nested]).to_csv(
        f"{args.output}/S6_nested_model_comparison.csv",
        index=False
    )
    pd.DataFrame([sens]).to_csv(
        f"{args.output}/S7_sensitivity_analysis.csv",
        index=False
    )
    pd.DataFrame([valid]).to_csv(
        f"{args.output}/S8_convergent_validity.csv",
        index=False
    )

    print(f"Analysis complete. Results saved to {args.output}")
```
