## Supplementary material for "Intellectual Property Literacy, Innovation Readiness and Innovation Practice in Syria’s Pharmaceutical Sector: A Cross-Sectional Study": README.docx

### **Innovation Capability Analysis Repository v1.0**

#### 📋 Overview

This repository contains the complete analytical code, de-identified dataset, and supplementary materials for a cross-sectional study of 303 pharmaceutical professionals examining the co-variation structure among intellectual property literacy, innovation attitudes, innovation readiness, and reported innovation practice within Syria's pharmaceutical sector.

**Study Design:** Cross-sectional survey
**Data Collection:** 10 March – 15 April 2026
**Ethics Approval:** Manara University (MU-URO-26-02)

![Reproducibility](<https://github.com/chadikhatib/innovation-capability-constraints/actions/workflows/reproducibility.yml/badge.svg>)

Tested with Python 3.12

#### 🗂️ Repository Structure

```
innovation-capability-constraints/
├── README.md                           ← This file
├── CITATION.cff                        ← Citation metadata
├── LICENSE                             ← MIT License
├── requirements.txt                    ← Python dependencies
│
├── data/
│   └── Deidentified_Dataset_N303.csv   ← De-identified dataset (N=303)
│
├── code/
│   └── Innovation_Capability_Analysis.py  ← Complete analysis script
│
├── supplementary/
│   ├── Table_S1_Participant_Flow.txt
│   ├── Table_S2_Demographics.txt
│   ├── Table_S3_Item_Level_Stats.txt
│   ├── Table_S4_Correlation_Matrix.txt
│   ├── Table_S5_ANOVA_PostHoc.txt
│   ├── Table_S6_Regression_Diagnostics.txt
│   ├── Table_S7_Sensitivity_Analyses.txt
│   ├── Table_S8_Convergent_Validity.txt
│
│
└── documentation/
    ├── SAP_STATISTICAL_ANALYSIS_PLAN_v1.0.txt
    ├── STROBE_CHECKLIST_v1.1.txt
    └── SUPPLEMENTARY_MATERIALS_INDEX.txt
```

#### 📊 Data Description

##### Dataset: Deidentified_Dataset_N303.csv

- **N = 303** pharmaceutical professionals
- **Format:** CSV (40 variables)
- **IDs:** Synthetic participant IDs (P001–P303)
- **De-identification:** All personal identifiers removed prior to analysis

##### Core Constructs

| Variable | Description | Type | Range | Items |
| --- | --- | --- | --- | --- |
| **PILS** | Pharmaceutical IP Literacy Score | Continuous | 0–100 | 9 binary (K2–K10) |
| **IAI** | IP Attitude Index | Continuous | 0–100 | 10 Likert (A1–A10) |
| **IRS** | Innovation Readiness Score | Continuous | 0–100 | 4 binary (R1–R4) |
| **IPI** | Innovation Practice Index | Continuous | 0–100 | 3 binary (P1–P3) |
| **Diagnostic_Gap** | IAI − IPI (descriptive) | Continuous | −40 to 94 | — |

##### Covariates

- Gender (Male/Female/Missing)
- Professional category (Pharmacist/Student/Academic)
- Experience level (<1, 1–5, 6–10, >10 years)

##### Missing Data Handling

- Gender non-response was coded as "2" in the raw dataset and recoded to missing (NA) during analysis
- No imputation performed
- Complete-case analysis for regression models
- One participant (0.3%) excluded from gender-stratified analyses only

#### 🔬 Measurement Model Statement

**All constructs are formative composite indicators, not reflective latent scales.**

- Inter-item homogeneity is **NOT** assumed.
- Internal consistency metrics (KR-20, Cronbach’s α) are reported for **transparency only.**
- These metrics are **NOT** used as validity criteria.
- Construct validity is evaluated via theoretical specification and empirical pattern coherence.
- Composite indicators were selected because the study was designed to characterize observable innovation-related capabilities rather than estimate latent psychological constructs.

*References: Coltman et al. (2008); Hanafiah (2020)*

#### 🚀 Quick Start

##### 1. Clone the Repository

git clone https://github.com/chadikhatib/innovation-capability-constraints.git
cd innovation-capability-constraints

##### 2. Install Dependencies

pip install -r requirements.txt

**Required packages:** - Python 3.12+ - pandas ≥ 2.0.0 - numpy ≥ 1.24.0 - scipy ≥ 1.10.0 - statsmodels ≥ 0.14.0 - matplotlib ≥ 3.7.0 - seaborn ≥ 0.12.0

##### 3. Run the Analysis

```
python code/Innovation_Capability_Analysis.py \
  --data data/Deidentified_Dataset_N303.csv \
  --output results/
```

##### 4. Outputs Generated

Running the script produces: - S2_descriptive_stats.csv - S4_correlation_matrix.csv - S5_specification_A.csv - S5_specification_B.csv

Note: These machine-readable outputs are intermediate analytical files and do not correspond directly to the formatted supplementary tables (Table S1–Table S8) included in the manuscript package.

#### 📈 Statistical Analysis Summary

| Analysis | Method | Key Output |
| --- | --- | --- |
| Descriptive statistics | Mean, SD, skewness, kurtosis | Table S2 |
| Correlations | Pearson r + Fisher z 95% CIs | Table S4 |
| Group comparisons | One-way ANOVA + Tukey HSD | Table S5 |
| Regression | OLS with HC3 robust SE | Table S6 |
| Nested comparison | ΔR², ΔAIC, ΔBIC + F-change | Table S6 |
| Diagnostics | VIF, Shapiro-Wilk, Breusch-Pagan, Durbin-Watson | Table S6 |
| Sensitivity | 5 alternative specifications | Table S7 |
| Convergent validity | Q10 vs PILS (Mann-Whitney U) | Table S8 |

#### ⚠️ Critical Alignment Statement

All analyses strictly follow **SAP v1.0**:

- ✅ No BCa bootstrap methods — Fisher z-transformation only
- ✅ No random seed dependency
- ✅ Nested model comparison: ΔR² + ΔAIC + ΔBIC framework
- ✅ All effect sizes follow predefined SAP hierarchy (η², f², Cohen’s d, r)
- ✅ All variables treated as formative composite indices (non-latent)

#### 📜 Ethics and Compliance

- **IRB Approval:** Manara University (MU-URO-26-02)
- **Declaration of Helsinki:** Fully adhered to
- **Informed Consent:** Obtained from all participants
- **Data Anonymization:** Fully applied prior to analysis
- **No competing interests declared**
- **No external funding reported**

#### 📚 Citation

If you use this repository, please cite:

```
@dataset{khatib2026ipinnovation,
  title={Innovation Capability under Institutional Constraints:
         De-identified Dataset and Analysis Code},
  author={Khatib, Chadi and Alkozy, Hala and Hamdan, Zainab and
          Isber, May and Mlhem, Jawa},
  year={2026},
  publisher={Zenodo},
  version={v2.3.4},
  doi={10.5281/zenodo.20716882},
  url={https://doi.org/10.5281/zenodo.20716882}
}
```

#### 📄 License

This project is licensed under the **MIT License** — see the LICENSE file for details.

#### 🔗 Links

- **GitHub Repository:** https://github.com/chadikhatib/innovation-capability-constraints
- **Zenodo Archive:** https://doi.org/10.5281/zenodo.20716882
- **Manuscript:** *[Submitted — peer review pending]*

#### 🙏 Acknowledgments

The authors gratefully acknowledge the institutional support and commitment to research excellence provided by the President of Manara University, the University Administration, and the Dean of the Faculty of Pharmacy.

The authors extend their sincere appreciation to the Syrian Pharmacists Syndicate, the Syrian Medical Syndicate, Latakia Hospital, Latakia University, and Al-Sham University for their logistical and professional support during the conduct of this study.

The authors are particularly grateful to the Syrian Association for Intellectual Property and Development (Mr. Yasser Saada) for their advisory contributions, and to Al-Manara Law Firm, Consultancies and Legal Studies, and specifically to General Manager and Legal Counsel Mr. Ahmad Talal Anis Karknawi for his expert legal guidance.

Finally, the authors wish to express their profound gratitude to all study volunteers and survey participants who generously devoted their time to complete the questionnaire on a voluntary basis. This research would not have been possible without their willingness to contribute.

The authors acknowledge the pharmacy students and academic staff of Manara University who participated in World Intellectual Property Day 2026 awareness activities conducted during the study period.

**Last Updated:** 2026-06-16 | **Version:** 1.0.0 | **Zenodo:** v2.3.4 | **DOI:** 10.5281/zenodo.20716882 | **Status:** Published
